# Efficacy of Mechanical Thrombectomy in Medium Vessel Occlusions of the MCA in the Extended Time Window

**DOI:** 10.1101/2024.06.16.24309009

**Authors:** Marcel Cedric Berger, Andreas Simgen, Philipp Dietrich, Weis Naziri

**Author notes:** Address correspondence to: Marcel Cedric Berger.

## Abstract

**Background:** Mechanical thrombectomy (MT) has significantly improved outcomes in acute ischemic stroke (AIS) due to large vessel occlusions (LVOs) up to 24 hours post-onset. The effectiveness of MT for medium vessel occlusions (MeVOs) in the M2 or M3 segments of the middle cerebral artery beyond 6 hours is less investigated.

**Methods:** This retrospective study analyzed 80 patients who underwent MT for primary, isolated M2 or M3 segment occlusions between January 2020 and August 2023. Patients were categorized by time from stroke onset to groin puncture into two groups: ≤6 hours (n=61) and >6 hours (n=19). Outcomes assessed included clinical severity (NIH Stroke Scale [NIHSS]), functional outcomes (modified Rankin Scale [mRS]), symptomatic intracranial hemorrhages (sICH), and reperfusion success (modified Thrombolysis in Cerebral Infarction [mTICI] scale).

**Results:** Mean onset-to-puncture time was 192±57 minutes for the ≤6 hours group and 611±327 minutes for the >6 hours group. Baseline NIHSS scores were 9.5 (IQR 9) and 7 (IQR 8), respectively (p=0.418). While the NIHSS improvement was greater in the ≤6 hours group (median: −5 vs. −2; p=0.028), both groups showed significant improvement from baseline NIHSS scores (p<0.001 and p=0.014). Rates of sICH were low in both groups (1.5% vs. 0.5%; p=0.421). Recanalization rates were lower in the >6 hours group (84.2% vs. 96.7%; p=0.084), with more attempts (2.37 vs. 1.66; p=0.024).

**Conclusion:** MT for M2 and M3 segment occlusions in the MCA shows benefits beyond 6 hours from stroke onset, with earlier treatment yielding greater improvement. Extending MT’s treatment window could be valuable for MeVOs in the MCA.

## Introduction

Acute ischemic stroke (AIS) continues to be a significant challenge in medical care, leading to considerable morbidity and mortality worldwide.^1^ The timing of intervention plays a crucial role in improving outcomes for patients, with earlier treatments generally leading to better neurological recoveries.^2^

Mechanical thrombectomy (MT) has proven effective for treating occlusions of the internal carotid artery and the M1 segment of the middle cerebral artery (MCA), up to 24 hours from onset in selected cases.^3,4^ However, the therapeutic approach for medium vessel occlusions (MeVOs) in the middle cerebral artery is less established. MeVOs, typically including segments such as M2/M3, A2/A3, and P2/P3, account for a substantial proportion of AIS cases (25-40%).^5^

Emerging evidence suggests potential benefits of MT in treating MeVOs, but current findings are primarily based on several small, non-randomized studies. Consequently, definitive recommendations are still pending from ongoing clinical trials, leaving some uncertainty in clinical decision-making, particularly regarding interventions beyond the conventional 6-hour treatment window.^6^

This paper seeks to contribute to the ongoing discussion by examining the safety and efficacy of mechanical thrombectomy in treating medium vessel occlusions, with a particular focus on cases treated within the extended time window (>6hrs).

## Methods

In this retrospective, single-center case series, we systematically reviewed consecutive patients with primary, isolated occlusions of the M2 or M3 segments of the middle cerebral artery, treated with mechanical thrombectomy at our institution from January 2020 to August 2023.

We stratified patients based on the interval from symptom onset to the commencement of intervention (onset to groin puncture time), dividing them into two cohorts: those treated within 6 hours (≤6 hours group) and those treated beyond 6 hours (>6 hours group). Wake-up strokes were included and categorized based on the last known well time. Strokes lacking proper documentation of onset were excluded from this analysis. All patients were followed up until discharge.

Mechanical thrombectomy eligibility followed institutional guidelines, primarily requiring a National Institutes of Health Stroke Scale (NIHSS) score of 4 or higher with a visualizable and accessible occlusion in the M2 or M3 segments. All patients underwent a routine imaging protocol, including unenhanced computed tomography (CT) of the brain, as well as CT angiography and CT perfusion imaging with the detection of salvageable tissue.

Patients displaying significant disabling symptoms—such as aphasia, hemianopia, or loss of hand function—were also considered candidates for MT, even with an NIHSS score below 4. High age, comorbidities, baseline functional status, and poor vascular anatomy were not considered rigid exclusion criteria for MT and were decided on a case-by-case basis.

Individuals presenting more than 24 hours post-symptom onset or lacking evidence of a potentially salvageable penumbra on perfusion imaging were excluded from MT.

For each patient, the collected data encompassed standard demographic information (age and sex), cerebrovascular risk factors (including diabetes, dyslipidemia, atrial fibrillation, and hypertension), as well as the clinical severity and functional disability at the time of admission and discharge. Additionally, mortality rates, the administration of recombinant tissue plasminogen activator (rtPA), time intervals, reperfusion outcomes, and rates of symptomatic intracranial hemorrhage (sICH) were documented.

Clinical severity and functional disability were evaluated using the NIHSS and the modified Rankin Scale (mRS) by a qualified neurologist at the time of admission and discharge. Additionally, the Alberta Stroke Program Early CT Score (ASPECTS) was utilized to assess initial ischemic changes, with scores derived from the admission CT scans. Reperfusion outcomes were assessed using the modified Thrombolysis in Cerebral Infarction (mTICI) scale, with scores determined by the operator at the end of the procedure. Successful recanalization was defined as mTICI ≥ 2b. The incidence of sICH was assessed with definitions adhering to those established in the ECASS-II trial.^7^

Routine CT imaging was performed within 24 hours post-procedure and subsequently according to clinical need. In cases of clinical deterioration, immediate CT imaging was undertaken.

All thrombectomies were conducted under general anesthesia, with the selection of the revascularization technique and device left to the discretion of the neurointerventionist.

All procedures adhered to the principles of the 1964 Helsinki Declaration and its subsequent amendments. The study received approval from the local ethics committee. Given the retrospective design of the study, the requirement for informed consent was waived.

### Statistics

Quantitative variables were presented as mean (± standard deviation) or median (interquartile range) and compared using either the Student’s t-test or Mann-Whitney U test. Categorical variables were expressed as numbers and percentages, and between-group comparisons were made using the chi-squared test or Fisher’s exact test as appropriate, with the latter used for cell frequencies less than 5. A significance level of p ≤ 0.05 was assumed. Statistical analysis was performed using SPSS v25 software (IBM Corp., Armonk, NY, USA).

## Results

In this study, a total of 80 patients with occlusions in the M2 or M3 segments of the middle cerebral artery were analyzed and divided into two cohorts based on the interval from symptom onset to intervention: those treated within 6 hours (≤6 hours group, n=61) and those treated after 6 hours (>6 hours group, n=19).

Both groups were comparable in terms of age, sex, and comorbidity prevalence (Table 1). As expected, the use of recombinant tissue plasminogen activator (rtPA) was notably higher in the ≤6 hours group (55.7% vs. 21.1%). In the >6 hours group, rtPA was administered within the usual time frame (≤4.5 hours), but the intervention began after more than 6 hours. Consequently, these patients were grouped accordingly.

**Table 1:**
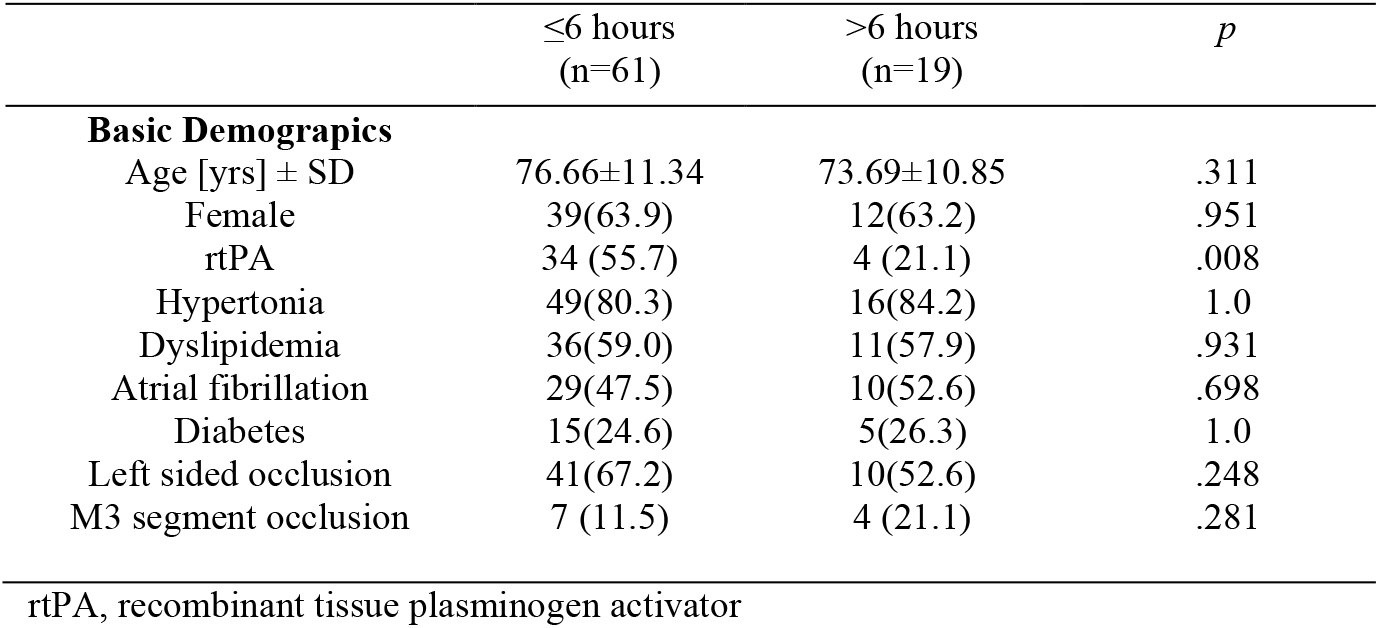
Baseline characteristics.

|  | ≤6 hours<br>(n=61) | >6 hours<br>(n=19) | *p* |
| --- | --- | --- | --- |
| **Basic Demographics** |  |  |  |
| Age [yrs] ± SD | 76.66±11.34 | 73.69±10.85 | .311 |
| Female | 39(63.9) | 12(63.2) | .951 |
| rtPA | 34 (55.7) | 4 (21.1) | .008 |
| Hypertonia | 49(80.3) | 16(84.2) | 1.0 |
| Dyslipidemia | 36(59.0) | 11(57.9) | .931 |
| Atrial fibrillation | 29(47.5) | 10(52.6) | .698 |
| Diabetes | 15(24.6) | 5(26.3) | 1.0 |
| Left sided occlusion | 41(67.2) | 10(52.6) | .248 |
| M3 segment occlusion | 7 (11.5) | 4 (21.1) | .281 |

rtPA, recombinant tissue plasminogen activator

**Table 2:**
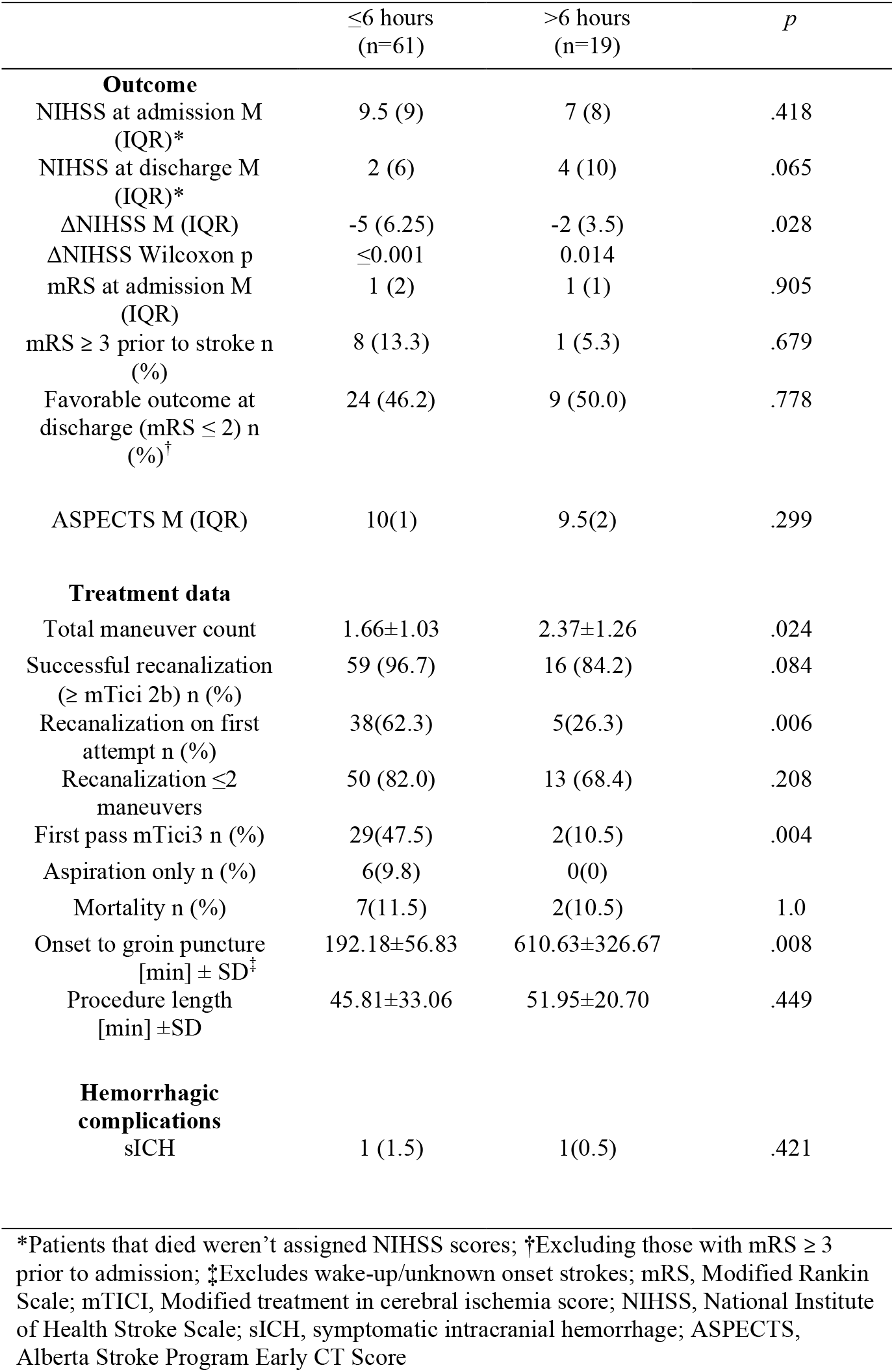
Treatment data, clinical outcomes, and complications.

The median Alberta Stroke Program Early CT Score (ASPECTS) showed no significant difference between the groups, with a median score of 10 (interquartile range [IQR]: 1) for the ≤6 hours group and 9.5 (IQR: 2) for the >6 hours group (p=0.299).

Upon admission, there was no significant difference in median NIHSS scores between the groups, with the ≤6 hours group at 9.5 (IQR: 9) and the >6 hours group at 7 (IQR: 8) (p=0.418). By discharge, the scores diverged, with the ≤6 hours group improving to a median score of 2 (IQR: 6), compared to 4 (IQR: 10) in the >6 hours group, approaching statistical significance (p=0.065).

Significant differences were noted in the change in NIHSS score (ΔNIHSS) from admission to discharge, indicating greater neurological improvement in the ≤6 hours group (median: −5, IQR: 6.25) compared to the >6 hours group (median: −2, IQR: 3.5; p=0.028). Both groups, however, showed significant improvement from their baseline scores (p<0.001 for the ≤6 hours group and p=0.014 for the >6 hours group).

No significant differences were observed in mRS scores at admission (median: 1, IQR: 2 for ≤6 hours group and median: 1, IQR: 1 for >6 hours group, p=0.905). The rate of favorable outcomes, defined as mRS ≤ 2 at discharge, was comparable between the groups (46.2% in the ≤6 hours group vs. 50% in the >6 hours group; p=0.778).

Treatment data revealed that the ≤6 hours group underwent fewer maneuvers on average (mean: 1.66 ± 1.03) compared to the >6 hours group (mean: 2.37 ± 1.26; p=0.024). The successful-recanalization rate (> mTICI 2b) was higher in the ≤6 hours group (96.7%) compared to the >6 hours group (84.2%), narrowly missing statistical significance (p=0.084).

The first-pass mTICI 3 rate, indicating a complete recanalization on the initial attempt, was significantly higher in the ≤6 hours group (47.5%) than in the >6 hours group (10.5%; p=0.004). Successful recanalization on first attempt also was significantly more often achieved in the ≤6 hours group (62.3%) compared to the >6 hours group (26.3%; p=0.006).

Regarding time metrics, the onset to groin puncture time was lower in the ≤6 hours group (mean: 192.18 ± 56.83 minutes) than in the >6 hours group (mean: 610.63 ± 326.67 minutes, p=0.008). Procedure length did not differ significantly between the groups, with the ≤6 hours group averaging 45.81 ± 33.06 minutes and the >6 hours group averaging 51.95 ± 20.70 minutes (p=0.449).

Hemorrhagic complications were low in both cohorts, with symptomatic intracranial hemorrhage occurring in 1.5% of the ≤6 hours group and 0.5% of the >6 hours group (p=0.421). Similarly, mortality rates were closely aligned, showing little variation: 11.5% in the ≤6 hours group compared to 10.5% in the >6 hours group (p=1.0).

## Discussion

This study provides preliminary insights into the efficacy of mechanical thrombectomy (MT) for treating medium vessel occlusions (MeVOs) within an extended time window. While landmark studies like DAWN and DEFUSE 3 have demonstrated MT’s effectiveness in proximal vessel occlusions up to 24 hours post-stroke, its application to MeVOs has been less explored.^3,4^

Emerging evidence suggests potential benefits of MT in MeVOs, yet international guidelines have not fully endorsed this treatment due to the scarcity of prospective studies. Ongoing clinical trials are expected to address this gap and may soon provide more concrete guidance on the application of MT in MeVOs, both within and beyond standard time frames.^6,8–10^

Our findings contribute to this evolving landscape by suggesting that the benefits of MT might extend to MeVOs not only in general but also specifically within the extended time window. This could influence clinical decision-making and shape future research directions.

In our cohort, we observed significant improvements in NIHSS scores from admission to discharge in both the ≤6 hours and >6 hours groups, though the extent of improvement varied with treatment timing. Specifically, the NIHSS scores improved from a median of 9.5 (IQR: 9) at admission to 2 (IQR: 6) at discharge in the ≤6 hours group, compared to an improvement from 7 (IQR: 8) to 4 (IQR: 10) in the >6 hours group. This difference underscores the benefits of early intervention, with the ≤6 hours group experiencing a median decrease in NIHSS of −5 (IQR: 6.25) versus −2 (IQR: 3.5) in the >6 hours group, indicating a more pronounced neurological recovery (p=0.028).

It is apparent from our results that the benefit of thrombectomy decreases as the time to recanalization increases. Patients who underwent MT later than 6 hours after symptom onset generally experienced smaller improvements, highlighting the importance of timely intervention. This trend is consistent with the well-established ‘time is brain’ principle, which posits that faster reperfusion translates to better neurological outcomes by minimizing ischemic brain injury.^2,11^

However, despite significant improvements in NIHSS scores in both groups, there was no significant difference in the rate of favorable outcomes at discharge (mRS≤2). Specifically, 46.2% of patients in the ≤6 hours group and 50.0% of patients in the >6 hours group achieved functional independence at discharge (p=0.778). These results are slightly lower than those reported in comparable studies, such as Bala et al., which showed functional independence rates at 90 days in late-window M2 occlusions of 59.6%. This discrepancy is likely due to the limited observation time in our study, which ended at patient discharge rather than following up to the 90-day mark.^9^

In our study, earlier intervention led to a more pronounced neurological recovery as indicated by NIHSS scores; however, this did not necessarily result in a higher proportion of patients achieving functional independence at discharge. Reasons for this might include the crudeness of the mRS, potentially overlooking subtle but significant differences in functional status. Additionally, mRS scores at discharge most likely don’t reflect the final outcome, as significant recovery and improvements can occur beyond the hospital stay and functional outcomes may diverge over time.

Furthermore, we observed a notable increase in the number of thrombectomy maneuvers in patients treated more than six hours after symptom onset, from 1.66 ± 1.03 in the ≤6 hours group to 2.37 ± 1.26 in the >6 hours group (p=0.024), accompanied by a significantly lower ‘first-pass mTICI 3’ rate (47.5% vs 10.5%; p=0.004). Additionally, there were notable differences in the rates of overall successful recanalization (≥ mTICI 2b), with a 96% success rate in those treated within six hours compared to 84.2% in those treated later, though this difference did not quite reach statistical significance (p=0.084). The first-pass successful recanalization rate was more than twice as high in the ≤6 hours group compared to the >6 hours group (62.3% vs 26.3%; p=0.006; Figure 1). These disparities may partially explain the observed differences in NIHSS scores, as studies examining large vessel occlusions have shown that both overall successful recanalization and first-pass recanalization are strongly associated with better clinical outcomes.^12,13^

**Figure 1:**
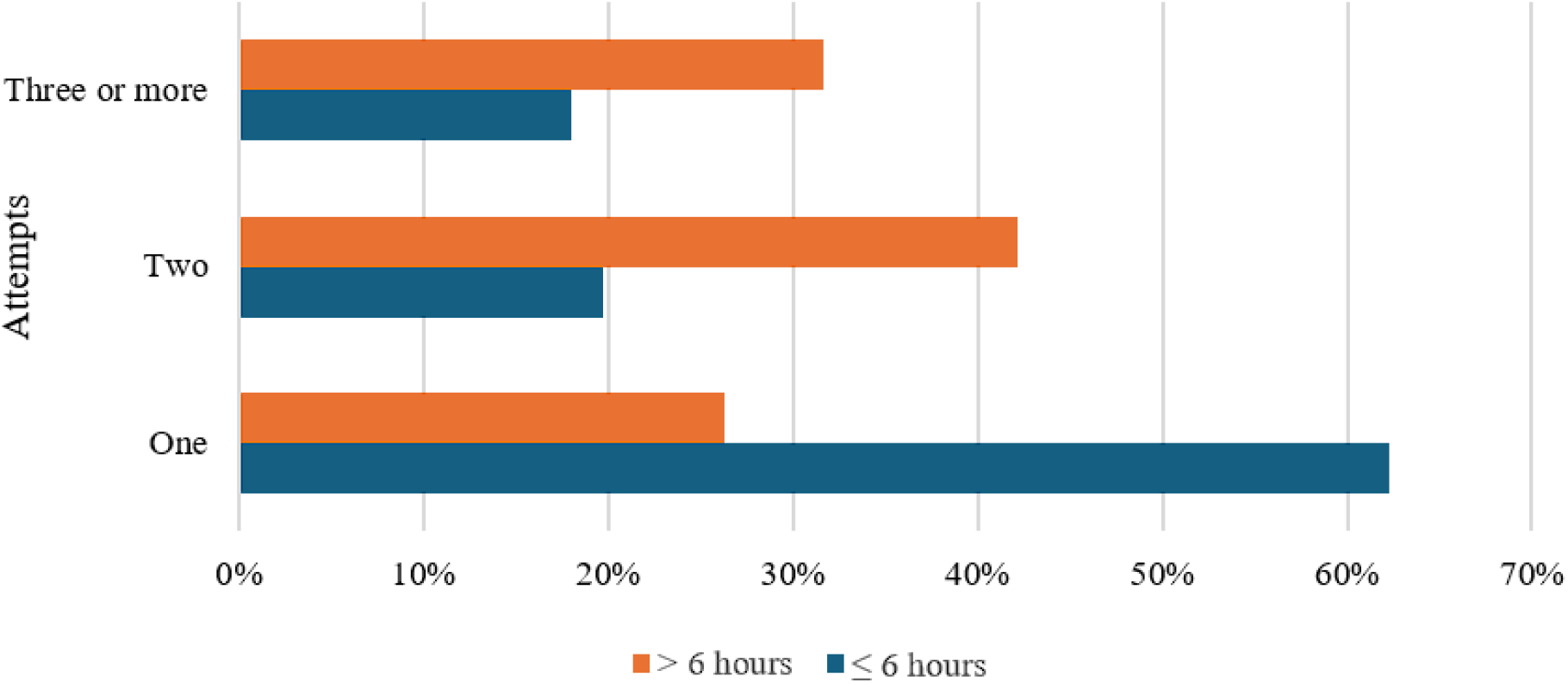
Distribution of total amount of endovascular maneuvers relative to treatment timing

The differences in maneuver count and recanalization rates might be due to changes in thrombus structure over time, which affect the technical success of MT. Thrombi that persist in vessels for extended periods tend to become more organized and harder due to increased fibrin cross-linking, making them more challenging to retrieve successfully.^14^

Interestingly, aspiration-only thrombectomies were exclusively observed in the ≤6-hour group. This finding may hint at the supposed greater efficacy of stent-retrievers over aspiration devices in older thrombi, as indicated in some studies, and may influence the choice of thrombectomy technique based on the timing of the intervention.^15,16^

It should be noted however that our study was not designed to evaluate the efficacy of different thrombectomy devices; the choice was at the neurointerventionist’s discretion, reflecting individual practitioner preference and experience. Further research seems warranted to better understand the efficacy and optimal use of different thrombectomy devices based on the timing of the intervention.

Despite the increased maneuver counts in the >6 hours group, we did not encounter an increase in symptomatic intracranial hemorrhage, as reported in some studies. The overall safety profile of MT remained favorable even when applied within an extended time window, with low rates of symptomatic intracranial hemorrhage in both cohorts (1.5% in the ≤6 hours group vs. 0.5% in the >6 hours group; p=0.421).^17^

Looking forward, it is crucial to refine patient selection criteria, potentially the choice of thrombectomy devices, and optimize timing strategies to maximize the benefits of mechanical thrombectomy for medium vessel occlusions. Insights from ongoing clinical trials will be instrumental in validating these findings and could broaden the application of mechanical thrombectomy to additional types of occlusions and clinical scenarios.

Given the study’s limitations, including its retrospective design, single-center scope, and small sample size, these results should be interpreted with caution. We cannot rule out the presence of selection bias in the patient selection process for thrombectomy, as it is possible that only patients with a high likelihood of benefit from endovascular thrombectomy, as judged by the treating physician, were treated. Notably, patients in the >6-hour group presented with an unusually high median ASPECTS score of 9.5, hinting at a slower evolution of ischemic damage. This factor could influence the generalizability of our results to the broader stroke population.

## Conclusion

In conclusion, our retrospective study confirms that mechanical thrombectomy is effective and safe for treating medium vessel occlusions in the M2 and M3 segments of the middle cerebral artery, even beyond the conventional 6-hour window. Our results reveal a trend towards improved outcomes with earlier interventions. Notably, the longer the delay before treatment, the more complex the intervention becomes, necessitating an increased number of maneuvers.

## Data Availability

The data that support the findings of this study are derived from patient records and contain sensitive information. As such, they are not publicly available due to privacy and ethical restrictions, including compliance with the General Data Protection Regulation (GDPR). De-identified data may be available from the corresponding author upon reasonable request.

